# Anticholinergic burden in middle and older age is associated with lower cognitive function, but not with brain atrophy

**DOI:** 10.1101/2022.09.04.22279576

**Authors:** J. Mur, R. E. Marioni, T. C. Russ, G. Muniz-Terrera, S. R. Cox

## Abstract

**Background:** Anticholinergic drugs block muscarinic receptors in the body. They are commonly prescribed for a variety of indications and their use has previously been associated with dementia and cognitive decline.

**Methods:** UK Biobank participants with linked health-care records (n=163,043, aged 40-71 at baseline), for about 17,000 of which MRI data was available, we calculated the total anticholinergic drug burden according to 15 different anticholinergic scales and due to different classes of drugs. We then used linear regression to explore the associations between anticholinergic burden and various measures of cognition and structural MRI, including general cognitive ability, 9 separate cognitive domains, brain atrophy, volumes of 68 cortical and 14 subcortical areas, and fractional anisotropy and median diffusivity of 25 white-matter tracts.

**Results:** Anticholinergic burden was modestly associated with poorer cognition across most anticholinergic scales and cognitive tests (7/9 FDR-adjusted significant associations, standardised betas (β) range: −0.039, −0.003). When using the anticholinergic scale exhibiting the strongest association with cognitive functions, anticholinergic burden due to only some classes of drugs exhibited negative associations with cognitive function, with β-lactam antibiotics (β=-0.035, p_FDR_<0.001) and opioids (β=-0.026, p_FDR_<0.001) exhibiting the strongest effects. Anticholinergic burden was not associated with any measure of brain macro- or microstructure (p_FDR_>0.08).

**Discussion:** Anticholinergic burden is weakly associated with poorer cognition, but there is little evidence for associations with brain structure. Future studies might focus more broadly on polypharmacy or more narrowly on distinct drug classes, instead of using purported anticholinergic action to study the effects of drugs on cognitive ability.

## Introduction

Anticholinergic drugs (anticholinergics) are medicines thought to block muscarinic receptors. Their anticholinergic action is ascertained by consulting anticholinergic scales that assign potency scores to individual drugs; the combined score for an individual patient is the anticholinergic burden (AChB). Anticholinergics are commonly prescribed for a variety of conditions^1^, and their transient side effects on cognition are well-known^2-6^. Moreover, their long-term use in old age^7^ and middle age^8-11^ has been associated with an increased risk of cognitive decline and dementia. It has been hypothesised^12^ that this relationship is due to central anticholinergic effects, affecting areas of the brain crucial for cognition^13-15^. Therefore, a relationship might exist between AChB, cognitive ability, and brain structure, even within the “normal” spectrum of cognitive functioning.

However, the existing evidence on this relationship is mixed. Most studies on anticholinergic prescribing in adults classify cognition as the absence vs. presence of a disorder or test separate cognitive modalities in isolation^16,17^. When measured this way, studies of AChB and cognitive ability often produce discordant results^16^. There are reports of positive associations between anticholinergic use and executive function^12,18-21^, associative learning^22^, visual-^23^, episodic-^24,25^, and short-term memory^26^, delayed and immediate recall^27^, language abilities^28^, visuospatial skills^28^, attention^28^, and reaction time^28^. However, some authors have found no evidence for delayed and immediate recall^21,22,28,29^, reaction time^22^, executive function^23,27^, language abilities^27,29^, working memory^25,27^, processing speed^25^, and implicit-^28^ and semantic^25^ memory. Additionally, because anticholinergic scales sometimes include different drugs and score the same drugs differently, they could represent another source of variation in reported findings.

It has been suggested that global composites of cognitive functioning might be more sensitive to subtle cognitive changes^16^. Individual test scores contain more random noise, and the results can limit generalisability and contribute to inconsistency among studies. On the other hand, general cognitive ability (sometimes referred to as general intelligence or *g*) represents shared variation across cognitive domains, is predictive of various social outcomes^30^, health outcomes^31,32^, mortality^33^, and is referenced in widely utilised diagnostic manuals^30^. Analysing large samples on multiple anticholinergic scales can further strengthen the reliability of the results.

Little is known about the neural correlates of the proposed anticholinergic-related cognitive decline. Associations exist between some brain measures, (e.g., white matter hyperintensities, total white- and grey matter volumes, and hippocampal volume) and cognitive ability (both individual domains and *g*) ^34-36^. To our knowledge, only three^8,12,37^ studies to date have assessed the relationship between these brain measures and regular anticholinergic use. While each study reported on associations between anticholinergic use and various metrics of brain structure and function, the sample size in each study was relatively small (n<800) and replication studies in larger samples are required. Furthermore, it is unclear if chronic anticholinergic use affects specific subcortical structures involved in cholinergic processing^38^ – the hippocampus, the putamen, and the amygdala. Finally, research is needed to probe potential differences between anticholinergic scales and between different classes of anticholinergics when exploring associations with cognitive function and cerebral correlates.

In our study – conducted using the UK Biobank – we calculated a latent factor of general cognitive ability (*g*) and utilised MRI-imaging measures and prescriptions linked from primary care, to study the association between AChB, *g*, and various brain structural MRI measures. Our goals were to assess (1) whether there existed differences between anticholinergic scales and (2) between drug classes in the association of AChB and cognitive ability, and (3) whether potential associations between AChB and cognitive ability were reflected in brain MRI measures, including brain atrophy, the volume of various cortical and subcortical brain structures, and measures of white matter microstructure.

## Methods

### Sample

UK Biobank^39^ is a prospective study whose participants were recruited between 2006 and 2010 when they were aged 37-73 years. During the initial assessment, demographic- and lifestyle questionnaires, physiological measurements, and cognitive tests were administered. A subset of participants later underwent MRI structural imaging and additional cognitive testing. For ∼230,000 participants, data on issued prescriptions and diagnoses are available. The diagnoses used were sourced from self-reported data, primary care, and secondary hospital care. Self-reported data were provided at the time of the assessments, while data from primary care and secondary hospital care are available until August 2017 and March 2021, respectively. Prescriptions are complete until May 2016 and were sourced from primary care. The prescription entries contained names and dates of drugs prescribed by general practitioners and the (mostly region-specific) suppliers of the prescription data. For the variables described below, we provide specific Field IDs (and links to the descriptions page for each field) in **Supplementary Table 1**.

### Cognitive ability

During the baseline assessment, most participants completed tests measuring visual declarative memory (Pairs Matching), processing speed (Reaction Time), with a subsample also completing tests of working memory (Numeric Memory), prospective memory (Prospective Memory), and verbal and numerical reasoning (Fluid Intelligence). During the imaging assessment, another subset of participants completed the above tests again, in addition to tests of executive function (Trail Making A and B, Tower Rearranging), verbal declarative memory (Paired Associate Learning), non-verbal reasoning (Matrix Pattern completion), crystallised ability (Picture Vocabulary), and another on processing speed (Symbol Digit Substitution) (**Supplementary Table 2**). Analyses of their psychometric properties in this sample have been reported previously^40,41^. We fitted a confirmatory factor analysis in a structural equation modelling (SEM) framework to calculate *g* from the cognitive tests (**Supplementary Figure 1 and Supplementary Table 3**), yielding two separate values, one for each assessment visit. SEM has been used to calculate *g* in UK Biobank before^35,42^; the proportional variance explained in our study is smaller (23% for the baseline assessment, 28% for the imaging assessment) than in prior work on in UK Biobank that used fewer cognitive tests^35^. For participants for whom this was possible, *g* from the imaging assessment was used in our analyses.

### Brain imaging

Since 2014, UK Biobank has been enhancing the dataset with imaging data that includes brain MRI^39,43^. It consists of imaging-derived phenotypes, whose acquisition and quality control have been previously described^44^. Briefly, brain imaging data were obtained at four data collection sites (Cheadle, Newcastle, Reading, and Bristol; all UK) using three identical scanners (3T Siemens Skyra), with a standard Siemens 32-channel receive head coil. Pre-processing and quality control were undertaken by the UK Biobank research team according to published procedures^44^. Our analyses included total brain volume, brain volumes of 68 cortical areas, 14 subcortical structures, functional anisotropy (FA), and mean diffusivity (MD) of 25 white matter tracts. The measures of brain volume were corrected for head size by multiplication with the T1-based scaling factor (UK Biobank field ID 25000). The brain regions and white-matter tracts used in the study are depicted in **Supplementary Figure 2**.

### Anticholinergic burden and drug classification

Anticholinergic scales typically score drugs on a 0-3 ordinal scale, with a higher score indicating greater anticholinergic potency. We considered 15 anticholinergic scales – thirteen^28,45-56^ were based on our previous analyses^1^ while two scales^57,58^ were identified through a recent review^7^ (**Supplementary Table 4**). Three scales^46,49,55^ were modified to include newer drugs^1,59^. One scale^51^ was modified so that drugs with “improbable anticholinergic action” were assigned an anticholinergic burden of 0.5 as was done before^1^. Using the British National Formulary (https://bnf.nice.org.uk/, last accessed on 11^th^ March 2021), we replaced brand names with generic names. Combination prescriptions containing several anticholinergics were each separated to yield multiple prescriptions, each containing a single anticholinergic. Each prescription was then assigned a potency score from each anticholinergic scale. For analysis, the cumulative AChB was calculated by summing the AChB-scores across all prescribed drugs in the sampling period. The sampling period excluded the year preceding the UK Biobank assessment to avoid acute effects of drugs. Prescriptions of drugs with ophthalmic, otic, nasal, or topical routes of administration were all assigned an anticholinergic score of 0, as previously reported^1,53-56^. Each drug was assigned to a class in the WHO Anatomical Therapeutic Chemical (ATC) Classification system (https://www.whocc.no, last accessed on 11^th^ March 2022) ^60^ that categorises drugs in a five-level hierarchy. In our analyses, the 1^st^ (anatomical main group) and 3^rd^ (pharmacological subgroup) levels were used.

### Data preparation

Prescriptions issued before the year 2000 and after the year 2015 were removed due to low ascertainment, and incomplete annual data, respectively^1^. Participants with a diagnosis of diseases that may affect brain structure or cognitive ability were removed. The data-cleaning process is depicted in **Supplementary Figure 3**. Outliers for numerical variables were defined as values lying four or more standard deviations or interquartile ranges beyond the mean or median, whichever was most appropriate according to the distribution. The total number of prescribed drugs and the AChB scores were strongly right-skewed due to the high numbers of zero values. For these variables, zeroes were removed before identifying outliers. All outliers were removed before analysis. Model assumptions were mostly met, but some models exhibited non-normality in the distribution of residuals (**Supplementary Figure 4**).

### Modelling

We applied principal component analysis to tract-specific FA and MD and used the first principal component to compute the “general” FA and MD (*g*FA and *g*MD), accounting for 44% and 50% of the variance, respectively. The standardised loadings and proportional variance for *g*FA and *g*FA are presented in **Supplementary Figure 5 and Supplementary Table 5**. We used linear regression models to estimate the association between AChB, cognitive ability, and brain structure. To compare anticholinergic scales, we modelled the association between *g* and AChB for each scale separately. This was later repeated for total brain volume as the outcome. The scale exhibiting the strongest association with *g* was selected for subsequent analyses. Second, we modelled the effects of AChB due to different drug classes on *g* and total brain volume. Finally, we computed the associations between AChB and the results from 9 cognitive tests, the volumes of 68 cortical areas, 14 subcortical areas, *g*FA and *g*MD, and FA and MD of 25 white-matter tracts. We also conducted two sensitivity analyses. First, we repeated the analyses on the association between AChB and *g* including only the year preceding the UK Biobank assessment to calculate AChB. Second, we computed the association between AChB according to each scale and *g*, while including an interaction term between AChB and age at assessment.

Each model was corrected for potential confounders, which included age at assessment, number of years over which the cumulative AChB was calculated, number of prescribed non-anticholinergics (different for each anticholinergic scale), data supplier of prescriptions (region-specific – two for England, and one each for Scotland and Wales), socioeconomic deprivation (higher values correspond to greater deprivation; range: −6.3-11.0) ^61^, smoking status (non-smoker, previous smoker, current smoker), frequency of alcohol consumption (daily or almost daily; three or four times a week; once or twice a week; once to three times a week; only on special occasions; never), level of physical activity (strenuous; moderate; mild) ^62^, BMI (kg/m^2^), *APOE*-carrier status, comorbidities count before the first assessment date (total number of distinct diagnoses codes), history of mood disorders, anxiety disorders, schizophrenia, diabetes, hypercholesterolemia, hypertension, and myocardial infarction before the assessment date. *APOE*-carrier status was defined through the *APOE* genotype, which is based on the nucleotides at SNP positions rs239358 and rs7412. Participants were denoted as ε2, ε3, or ε4 carriers, if they carried the ε2/ε2 or ε2/ε3 haplotype, ε3/ε3 or ε1/ε3 or ε2/ε4 haplotype, or ε3/ε4 or ε4/ε4 haplotype, respectively. Smoking status, alcohol consumption, physical activity, BMI, and genotype were ascertained at each of the two UK Biobank assessments; socioeconomic deprivation was ascertained during the baseline assessment.

When comparing anticholinergic scales, two additional models were run for which polypharmacy was the main predictor. The first of these models (P*olypharmacy* model) controlled for the same covariates as above, and the second (*Polypharmacy plus* model) further controlled for the total number of anticholinergics according to any scale. The models where a measure of brain imaging was the main outcome, were in addition to the covariates above controlled for age^2^, age*sex, age^2^*age, head position in the MRI-scanner (three coordinates), ethnicity, and assessment centre. The template for the linear models is described in **Supplementary Text 1**. Results are presented for models before the adjustment for polypharmacy and after adjustment for polypharmacy. Unless explicitly stated otherwise, the results refer to the fully adjusted models.

In analyses where a single anticholinergic scale was used (as opposed to comparing several scales), AChB was calculated using the scale by Durán et al. (2013) ^51^, as it exhibited the strongest association with *g* (see Results). All numerical variables were normalised to have a mean of 0 and a standard deviation of 1. When several independent models were run to predict the same outcome, p-values were corrected for multiple comparisons using the false discovery rate (FDR) ^63^. Otherwise, the p-threshold of 0.05 was used. Results are reported as standardised betas (β) and plotted with confidence intervals (CIs) adjusted for multiple comparisons (based on the Z-values of the quantile for the standard normal distribution for the FDR-adjusted p-values). All data cleaning and modelling were performed using R version 4.2.1 and Python version 3.9.7. The code is available at: https://github.com/JuM24/UKB-AChB-cognition-MRI.

## Results

### Sample

After removing outliers, among the 163,043 participants in our sample, ∼140,000 and ∼14,000 data points (depending on the model) were available for analyses of cognitive ability and brain imaging, respectively. The demographic- and lifestyle variables are presented in **Table 1**. While the imaging sample was older, the distribution of other variables was similar to the rest of the sample (**Supplementary Table 6**). In the period from 2000 to the year before the initial assessment, anticholinergics – depending on the anticholinergic scale – represented between 4.3% and 24.1% of prescriptions, with between 11.3% and 40.7% of participants prescribed an anticholinergic at least once (**Supplementary Table 7**). We have previously characterised anticholinergic prescribing and its longitudinal trends in UK Biobank in detail^1^.

**Table 1:** Demographic and lifestyle characteristics of the sample at the baseline assessment after the removal of outliers.

| Variable | Level | Median (IQR) or n (%) | N missing |
| --- | --- | --- | --- |
| Age |  | 58.3 (12.8) |  |
| Sex | Male | 72,184 (44.3) |  |
|  | Female | 90,859 (55.7) |  |
| Deprivation |  | -2.3 (3.9) | 174 |
| Alcohol consumption | Daily or almost daily | 32,244 (19.8) | 326 |
|  | Three or four times a week | 38,472 (23.6) |  |
|  | Once or twice a week | 43,328 (26.6) |  |
|  | Once to three times a month | 18,402 (11.3) |  |
|  | Only special occasions | 18,045 (11.1) |  |
|  | Never | 12,316 (7.6) |  |
| Smoking | Current smoker | 16,048 (9.9) | 772 |
|  | Previous smoker | 55,642 (34.3) |  |
|  | Non-smoker | 90,581 (55.8) |  |
| Physical activity | Strenuous | 16,531 (10.8) | 10,707 |
|  | Moderate | 97,776 (64.2) |  |
|  | Light | 38,029 (25.0) |  |
| BMI |  | 26.8 (5.8) | 1,029 |
| Data provider | England (Vision) | 14,393 (8.8) |  |
|  | Scotland | 9,571 (5.9) |  |
|  | England (TPP) | 122,120 (74.9) |  |
|  | Wales | 16,959 (10.4) |  |
| Mood disorder |  | 23.926 (14.7) |  |
| Anxiety disorder |  | 15.572 (9.6) |  |
| Schizophrenia |  | 590 (0.4) |  |
| Myocardial infarction |  | 7,335 (4.5) |  |
| Diabetes |  | 14,477 (8.9) |  |
| Hypercholesterolemia |  | 29,994 (18.4) |  |
| Hypertension |  | 54,124 (33.2) |  |
| Number of prior comorbidities |  | 86 (98) | 65 |
| Polypharmacy |  | 34 (96) |  |
| APOE carrier | ε2 | 20,549 (12.9) | 3,871 |
|  | ε3 | 98,084 (61.6) |  |
|  | ε4 | 40,539 (25.5) |  |

*Note:* Polypharmacy is the total number of prescriptions issued over the sampling period (differs among participants, range: 1-16 years). Sex, deprivation, alcohol consumption, smoking, physical activity, and BMI are self-reported or based on measurements during the baseline assessment. The variables are not scaled. BMI: body mass index; TPP: The Phoenix Partnership.

### AChB and cognition

When polypharmacy was not included as a control variable, all the tested anticholinergic scales exhibited significant negative associations with *g* (**Supplementary Table 8**). The scales by Durán et al. (2013) ^51^ and by Cancelli et al. (2008) ^48^ showed the strongest (β=-0.032, p^FDR^<0.001) and weakest (β =-0.009, p_FDR_<0.001) effects, respectively. When the models were additionally corrected for polypharmacy, the median effect size of AChB across scales was reduced by 31%, but associations of all anticholinergic scales except the scale by Cancelli et al. (2008) ^48^ (β=-4.4×10^−5^, p_FDR_=0.88) remained significant (**Figure 1A, Supplementary Table 8**). The scale by Durán et al. (2013) ^51^ retained the strongest association (β=-0.025, p_FDR_<0.001) (**Supplementary Table 9**). When the predictors were not standardised, this effect size corresponds to an at most 0.0017 decrease in *g* when AChB is increased by one standard deviation. The main predictors of each polypharmacy model also exhibited negative correlations with cognition, (P*olypharmacy*: β=-0.034 p_FDR_<0.001; P*olypharmacy plus:* β=-0.028, p_FDR_<0.001). The number of anticholinergics included in a scale was positively correlated with the strength of the observed effect when uncorrected for polypharmacy (r=0.70, p=0.004) and when corrected for polypharmacy (r=0.60, p=0.02, **Supplementary Figure 6**). I.e., the more drugs were identified as anticholinergic by an anticholinergic scale, the better predictor the scale was of lower *g*.

**Figure 1:**
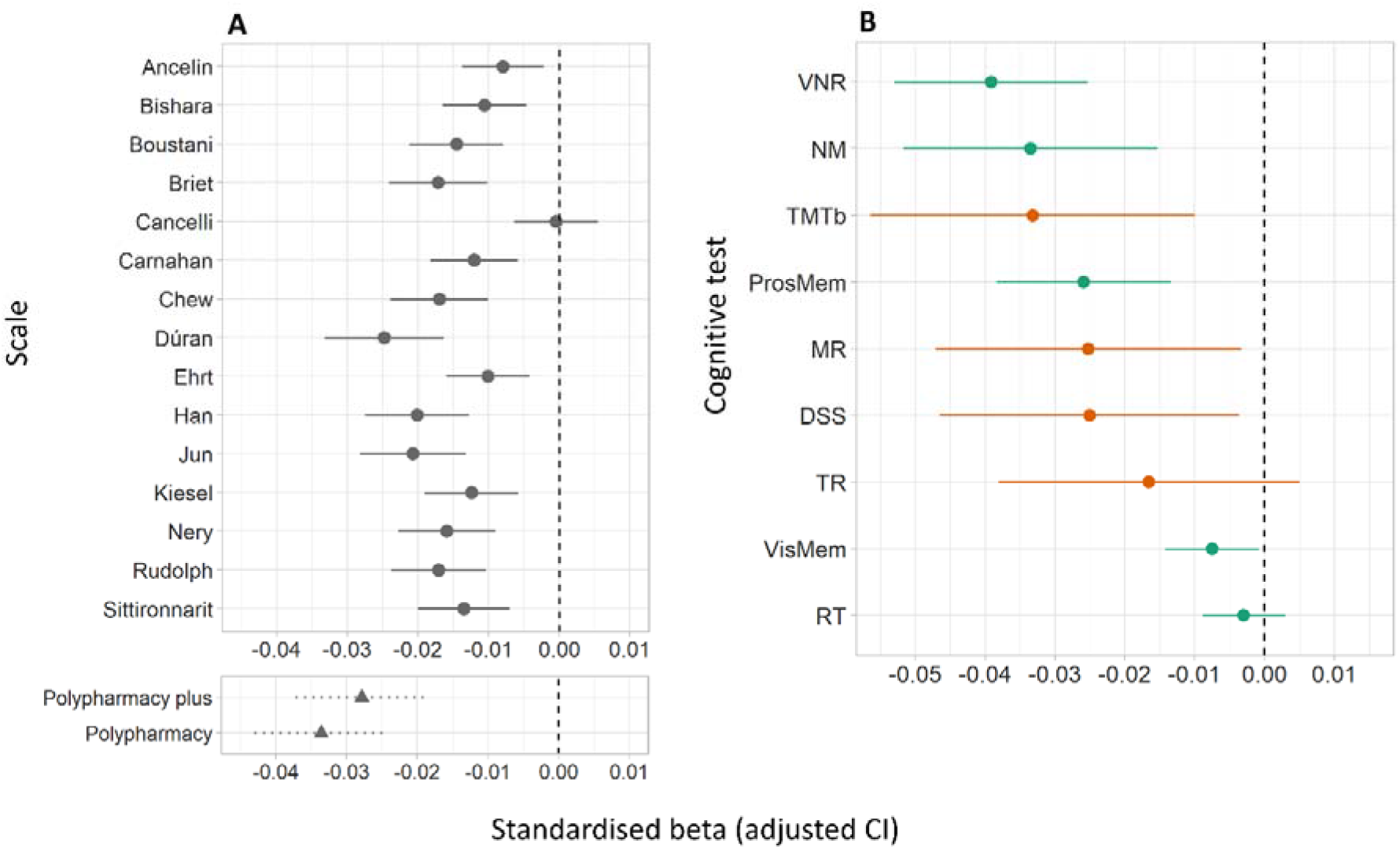
Associations between AChB and *g* for each anticholinergic scale **(A)** and associations between AChB according to the scale by Durán et al. (2013) ^51^ and each cognitive test included in the calculation of *g* **(B)**. Results are displayed as standardised βs. **A:** The y-axis indicates the main predictor for each model; in the upper panel, this was the AChB according to different anticholinergic scales; in the bottom panel, this was drug count (i.e., polypharmacy, controlled for in two different ways; see main text for details). **B:** The y-axis indicates the cognitive test used as the outcome. The colours refer to when the test was taken, with green indicating assessment at baseline and orange indicating assessment during the imaging visit. *Note*. DSS: digit Symbol Substitution test; TMTb: Trail-Making test; TR: Tower Rearranging test; ProsMem: Prospective Memory; VNR: Fluid Intelligence; MR: Matrix Pattern Completion; NM: Numeric Short-Term Memory; VisMem: Pairs-Matching test; RT: Reaction Time.

When a separate model was run for each cognitive test, AChB exhibited negative associations for each test. Among the cognitive tests, 7/9 were significant; Fluid Intelligence showed the strongest effect (β=-0.039, p_FDR_<0.01) and Reaction Time (β=-0.0030, p_FDR_=0.33) exhibited the weakest effects (**Figure 1B, Supplementary Table 10**).

When testing for the effects of drug classes, we found only limited instances in which higher AChB was associated with lower *g* (**Figure 2, Supplementary Table 11**). Among the pharmacological classes, AChB due to drugs for migraine (β=0.015, p_FDR_<0.001) showed positive associations with *g*. AChB due to most other drugs exhibited negative associations with *g*, with β-lactam antibiotics (β=-0.035, p_FDR_<0.001) and opioids (β=-0.026, p_FDR_<0.001) showing the strongest effects, corresponding to respectively 0.033 and 0.010 decreases in *g* for each increase of AChB by one standard deviation.

**Figure 2:**
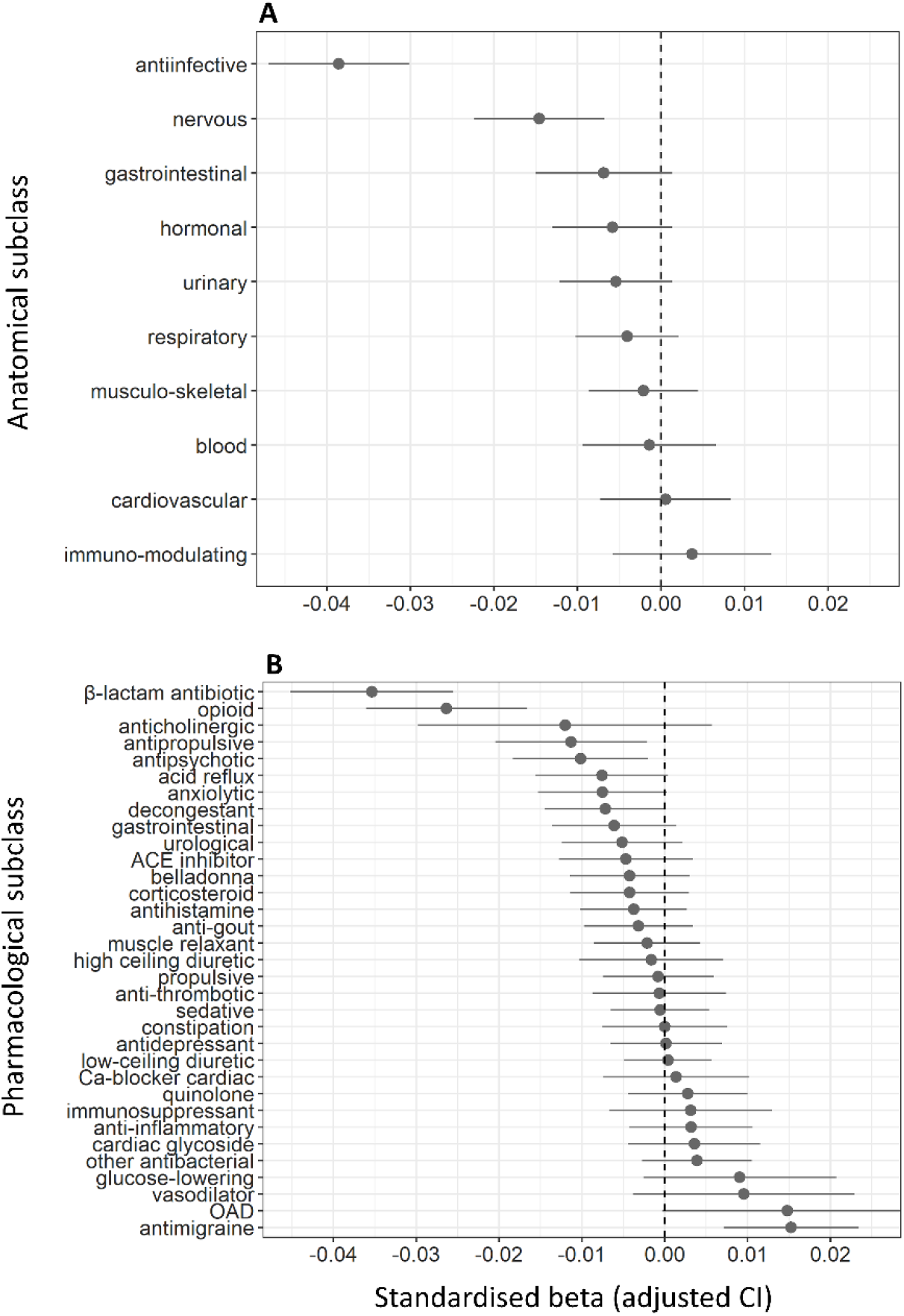
Associations between AChB according to the scale by Durán et al. (2013) ^51^ due to different classes of drugs on the one hand and *g* on the other. Results are displayed as standardised βs. **A:** classification of drugs based on anatomical class; **B:** classification of drugs based on pharmacological subclass. Classes prescribed to too few participants (<100) were not included in the models.

### ACB and brain-imaging measures

AChB was not associated with brain atrophy irrespective of the anticholinergic scale used (range of β=-0.004-0.017, p_FDR_≥0.21). While there were minor differences between the predictive power of different scales, the CIs overlapped across scale models and polypharmacy models (**Figure 3, Supplementary Tables 12, 13**).

**Figure 3:**
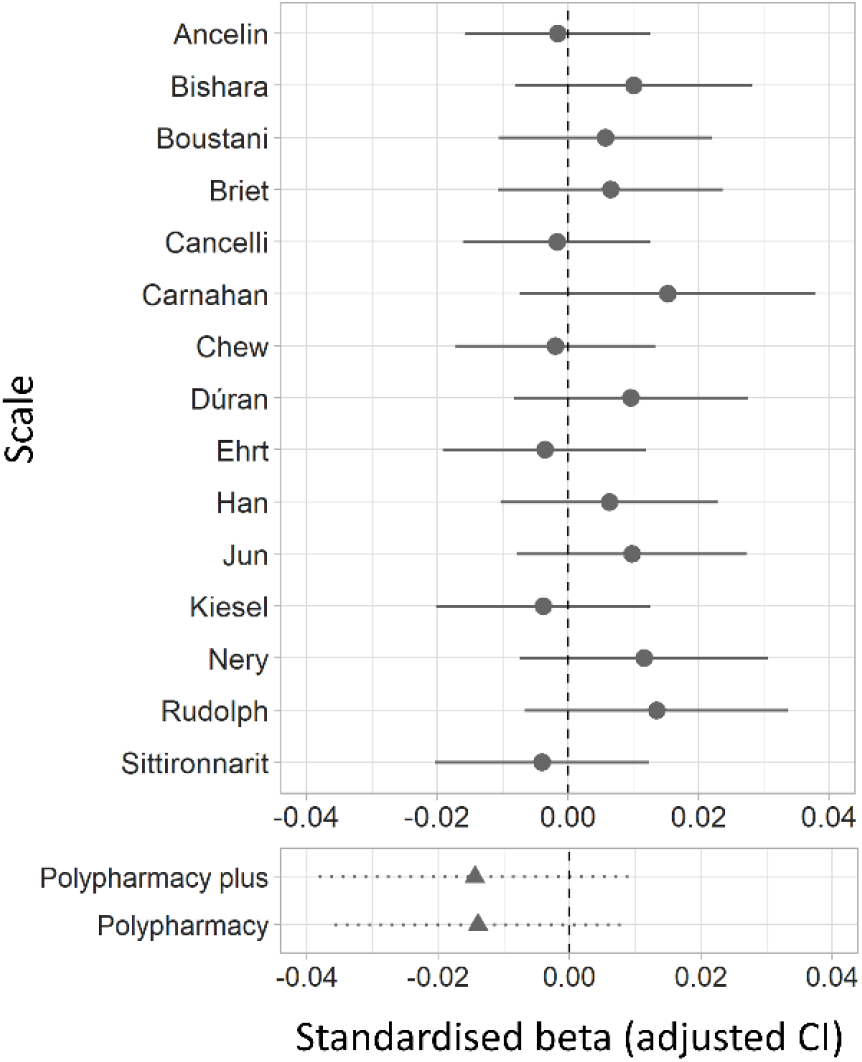
Associations between AChB and brain atrophy for each anticholinergic scale. The y-axis indicates the main predictor for each model; in the upper panel, this was the AChB according to different anticholinergic scales; in the bottom panel, this was drug count (i.e., polypharmacy, adjusted for covariates in two different ways; see main text for details). Results are displayed as standardised βs.

AChB was also not associated with the volume of any cortical (range of β=-0.018-0.028, p_FDR_≥0.26) or subcortical (β range=-0.007-0.024, p_FDR_≥0.08) brain region, or the microstructure of white matter tracts (range of β=-0.015-0.014, all p_FDR=_0.98) (**Supplementary Table 14**). When exploring associations of AChB due to different classes of drugs with brain atrophy, the effect sizes were again small, with most CIs overlapping with zero (**Supplementary Table 15, Figure 4**). AChB due to no single drug class was associated with brain atrophy.

**Figure 4:**
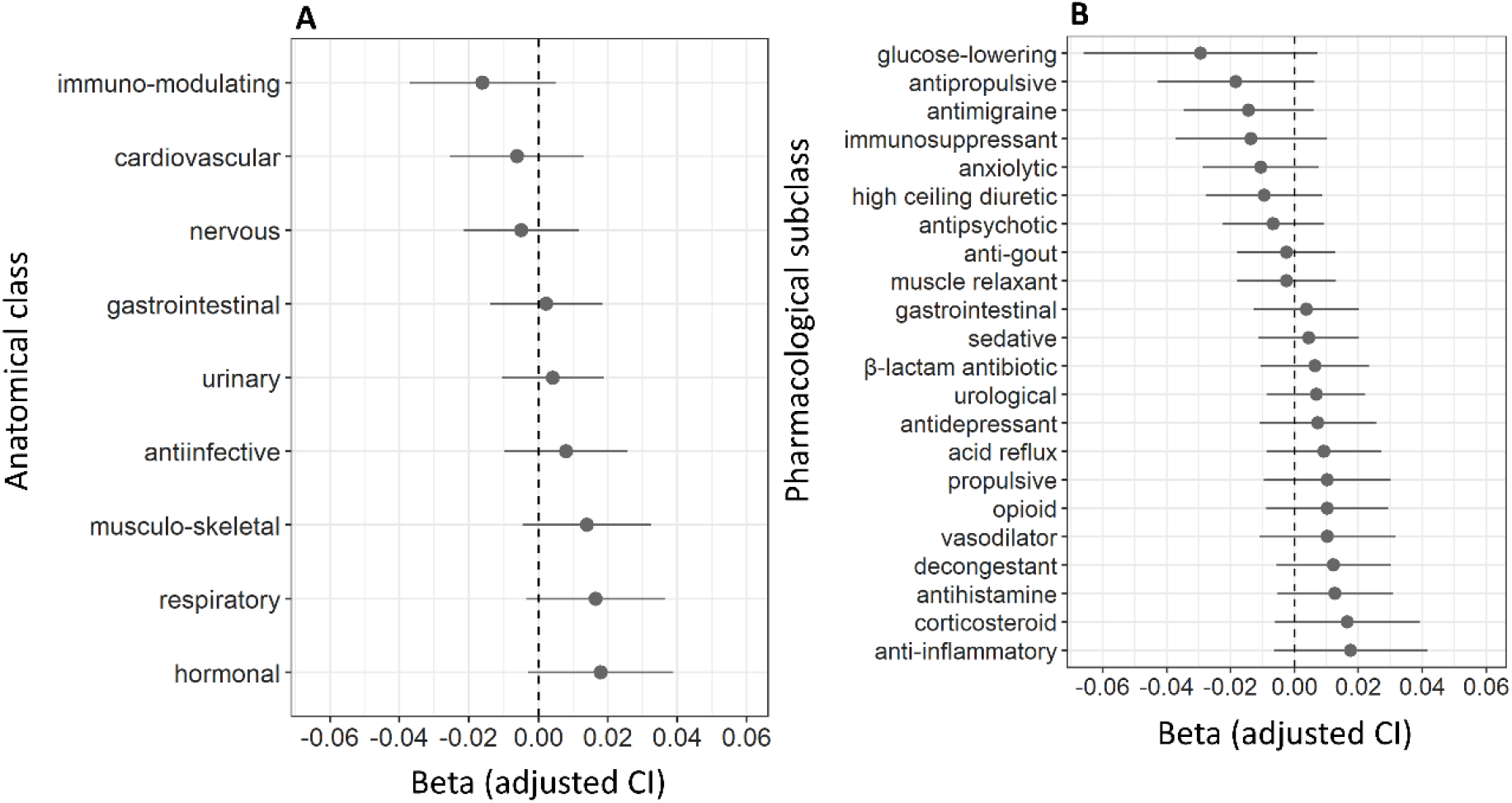
Associations between AChB according to the scale by Durán et al. (2013) ^51^ calculated for the year before the assessment due to different classes of drugs on the one hand and total brain volume on the other. Results are displayed as standardised βs. **A:** classification of drugs based on anatomical class; **B:** classification of drugs based on pharmacological subclass. Classes with too few prescriptions in the sample (<100) were not included in the models.

### Sensitivity analyses

When the analyses on the associations between AChB and cognitive function were repeated using only AChB in the year before the assessment as the predictor (**Supplementary Tables 16-19**), the results exhibited similar trends to those observed in the main analyses. Most anticholinergic scales tended to negatively associate with cognitive function, albeit the effect sizes were smaller.

Additionally, AChB was associated with lower performance in 1/5 cognitive tests available for this analysis. Furthermore, AChB due to β-lactam antibiotics and opioids again exhibited the strongest negative associations with *g*.

When *g* was modelled with the inclusion of an interaction term between age at assessment and AChB, the interaction was not significant (β=3.0×10^−4^, p=0.38), indicating that the observed effect sizes were not substantially larger or smaller in older compared to younger participants.

## Discussion

In this study, we found that most of the 15 studied anticholinergic scales exhibited significant associations with cognitive ability. This remained the case after controlling for multiple potential confounds, including the history of certain disorders and polypharmacy. Interestingly, the size of the effect was not moderated by age – middle-aged and older adults showed similar AChB-cognitive associations. While the positive association between higher AChB and lower cognitive ability largely agrees with previous studies on the topic, past results have been mixed^7,16^. One potential source of heterogeneity between studies is different control for polypharmacy, which may alter the results considerably. In our study, the addition of polypharmacy substantially decreased the size of the observed effects and was a stronger predictor of lower cognitive ability than AChB according to any of the studied anticholinergic scales. Another source of heterogeneity may be the differential effect of distinct drug classes. We found large differences between drug classes when predicting cognitive ability, with β-lactam antibiotics exhibiting larger effects than other drug classes. Moreover, antimigraine drugs were associated with higher cognitive ability. The effect of a general anticholinergic score may thus strongly depend on the structure of the sample and the precise prescribing characteristics of the participants.

In our study, general AChB was not predictive of any measure of brain structural MRI studied, including the volumes of 68 cortical and 14 subcortical areas, and measures of brain microstructure for 25 white-matter tracts. These findings are in contrast with previous research. To our knowledge, three studies have explored the association between anticholinergic use and brain structure. They found anticholinergic use to associate with reduced cortical volume and reduced temporal lobe thickness^12^, increased rates of brain atrophy^8^ and reduced grey matter density and functional connectivity in the nucleus basalis of Meynert^64^.

It is unclear why our results from MRI structural imaging diverge from previous findings, as the studies described above display a range of characteristics that overlap with our own, including longitudinal data^8^, control for polypharmacy^12^, and the inclusion of middle-aged participants^8^. One possibility is that the previous studies mostly classified the predictor (e.g., anticholinergic users vs. non-users), while we used a continuous measure of AChB. The pitfalls of categorisation and the loss of power for true effects have been discussed before^65^. Furthermore, the size of our imaging sample (∼17,000) was more than an order of magnitude greater than in previous studies (<800). As has been recently reported^66^, brain-wide association studies may require thousands of participants to minimise effect size inflation and increase replication rates. Finally, the above studies focused on cognitive disorders or decline later in life, with one reporting an effect for specifically those participants that later developed mild cognitive impairment^64^. It is possible that while brain atrophy occurs in ageing or dementia, subtle cellular changes in the cholinergic system occur before that but are not measurable by structural and diffusion MRI. This could include changes in the proportions or the integrity of muscarinic receptor subtypes or a shift in the balance of oscillation frequencies of neural networks.

Our study exhibits several advantages, including the use of a far larger sample than ever before in this area, use of linked prescriptions from primary care across a long period, exploration of several outcomes, the use of a latent factor of cognitive ability, and the comparison of different anticholinergic scales and classes of drugs. Furthermore, our models carefully incorporated several important control variables, including the history of relevant disorders, polypharmacy, and several lifestyle and demographic factors. Finally, we adopted a robust approach to measuring cognitive ability that can reduce variability common in the assessment of separate cognitive domains.

However, we recognise several limitations. First, the UK Biobank sample is on average less deprived and healthier than the UK population^67^ and thus not representative. Participants in the imaging subsample exhibit even better indicators of psychological and physical health than the UK Biobank average. Both factors likely result in an underestimate of the effects present in the population. Second, the prescriptions included in our study do not incorporate over-the-counter drugs and we also have no information on how many prescriptions were dispensed or taken by participants. Third, brain imaging was sometimes performed after the coverage for prescriptions had concluded and the drugs potentially prescribed in the intervening period were not accounted for. This likely decreased the accuracy of our AChB measure for those participants. Fourth, our study was cross-sectional and did not assess longitudinal changes in cognitive function and brain structure. This prevented us from establishing the sequence of events and from assessing associations between anticholinergic use and within-person changes. Finally, because AChB correlates with the number of anticholinergics, the effects of polypharmacy due to the use concurrently of several anticholinergic drugs and intrinsic anticholinergic activity of those drugs could not be completely separated.

Both the present study, as well as previous analyses have reported polypharmacy more broadly to be associated with poorer cognitive ability^68,69^ and dementia^70^. A recent medication-wide association study^71^ found that among 744 medicines, 30% were associated with dementia. Additionally, previous studies have reported on differences between drug classes in the association between AChB and dementia^9-11^. This finding was extended in the present study of general cognitive ability in a non-pathological sample. These results support a more nuanced approach that distinguishes between different classes of drugs beyond their assumed anticholinergic effects. For drug classes for which associations with lower cognitive ability or dementia can be demonstrated, more studies are needed to determine the effects of chronic use earlier in life, the impact of discontinuation, and the potential neural correlates.

While the effect sizes observed in our study were modest, for complex, multicausal outcomes – especially in a large and relatively healthy sample – this is to be expected. For example, ACE inhibitors – one of the most common drugs to treat hypertension – have been shown to reduce systolic/diastolic pressure by merely −8/-5 mm Hg^72^. When considered in the long-term and on the scale of entire populations, even tiny effects can accumulate to produce substantial health- and economic consequences for society. Given sufficient confidence in a drug-outcome relationship and the availability of alternative treatments, changes in prescribing represent an intervention that is relatively simple to implement. This should serve as additional motivation for further research in the field.

## Data Availability

All data produced in the present study are available upon request from UK Biobank.

https://github.com/JuM24/UKB-AChB-cognition-MRI

## Acknowledgements

This research was funded in whole, or in part, by the Wellcome Trust [108890/Z/15/Z]. For the purpose of open access, the author has applied a Creative Commons Attribution (CC BY) licence to any Author Accepted Manuscript version arising from this submission. REM is supported by Alzheimer’s Research UK major project grant ARUK-PG2017B-10 and by an Alzheimer’s Society major project grant AS-PG-19b-010. JM is supported by funding from the Wellcome Trust 4-year PhD in Translational Neuroscience—training the next generation of basic neuroscientists to embrace clinical research [108890/Z/15/Z]. JM and TCR are members of the Alzheimer Scotland Dementia Research Centre funded by Alzheimer Scotland. TCR is employed by NHS Lothian and the Scottish Government and has received grants from the Royal Society of Edinburgh [1689], the Scottish Government, the Chief Scientist Office [NRS NDN and CGA/19/05], the UK National Health Service, and UK Research and Innovation [ES/W001349/1]. SRC is supported by a Sir Henry Dale Fellowship, jointly funded by the Wellcome Trust and the Royal Society [221890/Z/20/Z] and has received grants from Age UK [The Disconnected Mind project], the UK Medical Research Council [MR/R024065/1], the National Institutes of Health [R01AG054628; 1RF1AG073593] and the UK Biotechnology and Biological Sciences Research Council and the Economic and Social Research Council (BB/W008793/1; ES/T003669/1). GMT has received funding from the Medical Research Council Methodology Board. The authors thank all participants of the UK Biobank for providing data for the study, and Dr Michelle Luciano (Department of Psychology, University of Edinburgh) for managing UK Biobank data application 10279.

## Conflicts of interest

REM has received consulting fees from the Epigenetic Clock Development Foundation and speaker fees from Illumina. TCR has received fees for medicolegal work from private solicitors. SRC has received speaker fees from the Society of Biological Psychiatry. GMT has received consulting fees for grants funded by the NIH. JM has nothing to disclose.

## Notes

### Author Declarations

The Research Ethics Committee (REC) of the Univeristy of Edinburgh granted ethical approval for the study (reference 11/NW/0382) and the current analysis was conducted under data application 10279.

### Summary of Updates

Previous version stated that AChB according to "most" drug classes was associated with lower general cognitive ability, although only some demonstrated significant associations. This has now been corrected. Also, "reduced" in the title has been altered to "lower" so as not to suggest repeated/longitudinal measurements.

