## Supplemental tables and figures for "Anticholinergic burden in middle and older age is associated with lower cognitive function, but not with brain atrophy": supplement.docx

**Supplementary Table 1:** UK Biobank variables and their Field IDs used in the study.

| **Variable** | **Field ID** | **UK Biobank showcase link** |
| --- | --- | --- |
| Demographic and lifestyle | | |
| Age | 21003 | <https://biobank.ndph.ox.ac.uk/showcase/field.cgi?id=21003> |
| Sex | 31 | <https://biobank.ndph.ox.ac.uk/showcase/field.cgi?id=31> |
| Deprivation | 189 | <https://biobank.ndph.ox.ac.uk/showcase/field.cgi?id=189> |
| Alcohol consumption | 1558 | <https://biobank.ndph.ox.ac.uk/showcase/field.cgi?id=1558> |
| Smoking | 20116 | <https://biobank.ndph.ox.ac.uk/showcase/field.cgi?id=20116> |
| Physical activity | 6164 | <https://biobank.ndph.ox.ac.uk/showcase/field.cgi?id=6164> |
| BMI | 21001 | <https://biobank.ndph.ox.ac.uk/showcase/field.cgi?id=21001> |
| Data provider | 42039 | <https://biobank.ndph.ox.ac.uk/showcase/field.cgi?id=42039> |
| *APOE* carrier | 22418 | <https://biobank.ndph.ox.ac.uk/showcase/field.cgi?id=22418> |
| Ethnicity | 21000 | <https://biobank.ndph.ox.ac.uk/showcase/field.cgi?id=21000> |
| Imaging assessment centre | 54 | <https://biobank.ndph.ox.ac.uk/showcase/field.cgi?id=54> |
| Date of attending assessment centre | 53 | <https://biobank.ndph.ox.ac.uk/showcase/field.cgi?id=53> |
| Prescriptions and diagnoses | | |
| Inpatient diagnoses | 41270 41271 41280 41281 | <https://biobank.ndph.ox.ac.uk/showcase/field.cgi?id=41270> <https://biobank.ndph.ox.ac.uk/showcase/field.cgi?id=41271> <https://biobank.ndph.ox.ac.uk/showcase/field.cgi?id=41280> <https://biobank.ndph.ox.ac.uk/showcase/field.cgi?id=41281> |
| Primary care diagnoses | 42040 | <https://biobank.ndph.ox.ac.uk/showcase/field.cgi?id=42040> |
| Primary care prescriptions | 42039 | <https://biobank.ndph.ox.ac.uk/showcase/field.cgi?id=42039> |
| Self-reported illness | 20002 | <https://biobank.ndph.ox.ac.uk/showcase/field.cgi?id=20002> |
| Cognitive tests | | |
| DSS | 23324 | <https://biobank.ndph.ox.ac.uk/showcase/field.cgi?id=23324> |
| MR | 6373 | <https://biobank.ndph.ox.ac.uk/showcase/field.cgi?id=6373> |
| NM | 4282 | <https://biobank.ndph.ox.ac.uk/showcase/field.cgi?id=4282> |
| ProsMem | 20018 | <https://biobank.ndph.ox.ac.uk/showcase/field.cgi?id=20018> |
| RT | 20023 | <https://biobank.ndph.ox.ac.uk/showcase/field.cgi?id=20023> |
| TMTb | 6350 | <https://biobank.ndph.ox.ac.uk/showcase/field.cgi?id=6350> |
| TR | 21004 | <https://biobank.ndph.ox.ac.uk/showcase/field.cgi?id=21004> |
| VisMem | 399 | <https://biobank.ndph.ox.ac.uk/showcase/field.cgi?id=399> |
| VNR | 20016 | <https://biobank.ndph.ox.ac.uk/showcase/field.cgi?id=20016> |
| MRI imaging | | |
| Total brain volume | 25010 | <https://biobank.ndph.ox.ac.uk/showcase/field.cgi?id=25010> |
| Cortical areas volume | 27205-27235 27298-27328 | <https://biobank.ndph.ox.ac.uk/showcase/label.cgi?id=196> |
| Subcortical areas volume | 25011-25024 | <https://biobank.ndph.ox.ac.uk/showcase/label.cgi?id=1102> |
| FA and MD of white matter | 25488-25514 25515-25541 | <https://biobank.ndph.ox.ac.uk/showcase/label.cgi?id=135> |
| Head position in scanner | 25756-25758 | <https://biobank.ndph.ox.ac.uk/showcase/field.cgi?id=25756>  <https://biobank.ndph.ox.ac.uk/showcase/field.cgi?id=25757> <https://biobank.ndph.ox.ac.uk/showcase/field.cgi?id=25758> |
| T1 head size scaling factor | 25000 | <https://biobank.ndph.ox.ac.uk/showcase/field.cgi?id=25000> |

**Supplementary Table 2**: All cognitive tests available in UK Biobank, with the mean and standard deviation for each test (before scaling), and the numbers of participants from our sample (and % of the sample) that underwent testing at either the baseline or the imaging assessment. The greyed-out tests were not used in our study, either due to measuring crystallised cognitive ability (Picture Vocabulary) or due to ceiling-effects (Paired Associative Learning). The Trail Making Tests, Tower Rearranging, Matrix Pattern Completion, and Symbol Digit Substitution were not administered during the baseline assessment. The data has been cleaned for outliers, defined as values four or more standard deviations below or above the mean.

| **Cognitive test** | **Baseline assessment** | | | **Imaging assessment** | | |
| --- | --- | --- | --- | --- | --- | --- |
|  | **N (%)** | **Mean** | **Std.** | **N (%)** | **Mean** | **Std.** |
| Pairs Matching | 161,944 (99.3) | 1.42 | 0.64 | 18,649 (11.4) | 1.34 | 0.62 |
| Reaction Time | 160,571 (98.5) | 6.30 | 0.18 | 18,546 (11.4) | 6.37 | 0.17 |
| Numeric Memory | 25,565  (15.7) | 6.71 | 1.32 | 13,856 (8.5) | 6.78 | 1.27 |
| Prospective Memory* | 63,623  (39.0) | 14,493 (22.7) |  | 18,746 (11.5) | 3,133 (16.7) |  |
| Fluid Intelligence | 61,599  (37.8) | 6.0 | 2.13 | 18,403 (11.3) | 6.60 | 2.06 |
| Trail Making |  |  |  | 13,082 (8.0) | 6.29 | 0.34 |
| Tower Rearranging |  |  |  | 13,421 (8.2) | 9.86 | 3.25 |
| Paired Associative Learning |  |  |  |  |  |  |
| Matrix Pattern Completion |  |  |  | 13,539 (8.3) | 7.93 | 2.13 |
| Picture Vocabulary |  |  |  |  |  |  |
| Symbol Digit Substitution |  |  |  | 13,543 (8.3) | 18.8 | 5.29 |

**Note:* Prospective memory was a binary variable; the values indicate the numbers (and %) of participants with correct recall.
**Supplementary Figure 1:** Path diagram for SEM calculating the latent *g* from individual cognitive tests administered during the baseline assessment (**top**) or during the imaging assessment (**bottom**). The arrows depict standardised loadings (the latent variable and the observed variables have a variance of 1), with positive loadings depicted with grey one-way arrows and negative loadings depicted with red one-way arrows. The dotted line indicates that the (unstandardised) loading was fixed to 1. MR and VNR on the one hand, and DSS and RT one the other hand measure similar cognitive abilities and residual correlations between them were included in the model; this is depicted by two-way arrows between the cognitive tests. Two-way arrows within individual cognitive tests represent residual variances. The total variance in cognitive test scores explained by the latent factor was 0.23 for the baseline assessment and 0.28 for the imaging assessment. The model-fit statistics are displayed on the right side of each diagram.

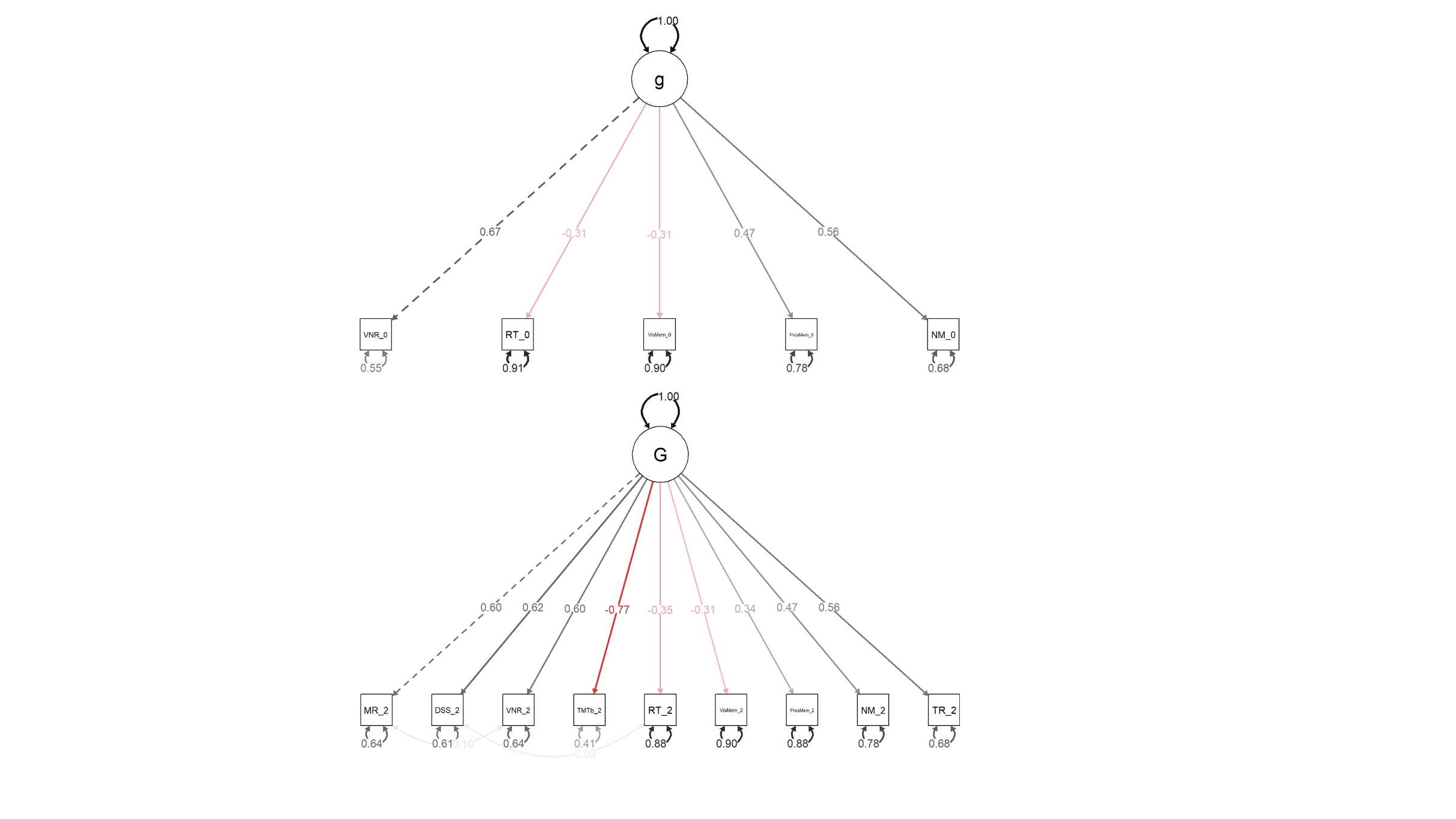

RMSEA: 0.022
SRMR: 0.021
CFI: 0.974
TLI: 0.948

RMSEA: 0.041
SRMR: 0.023
CFI: 0.965
TLI: 0.950

*Note*: MR: Matrix Pattern Completion; DSS: Digit Symbol Substitution; VNR: Fluid Intelligence; TMTb: Trail Making Test B; RT: Reaction Time; VisMem: Pairs Matching; ProsMem: Prospective Memory; NM: Numeric Memory; TR: Tower Rearranging.

**Supplementary Figure 2**: Cortical regions from the Desikan-Killiany neuroanatomical atlas (**top**), white matter tracts (**bottom left**) and subcortical structures (**bottom right**) measured in the present study. Figures reused from previous studies^1,2^.

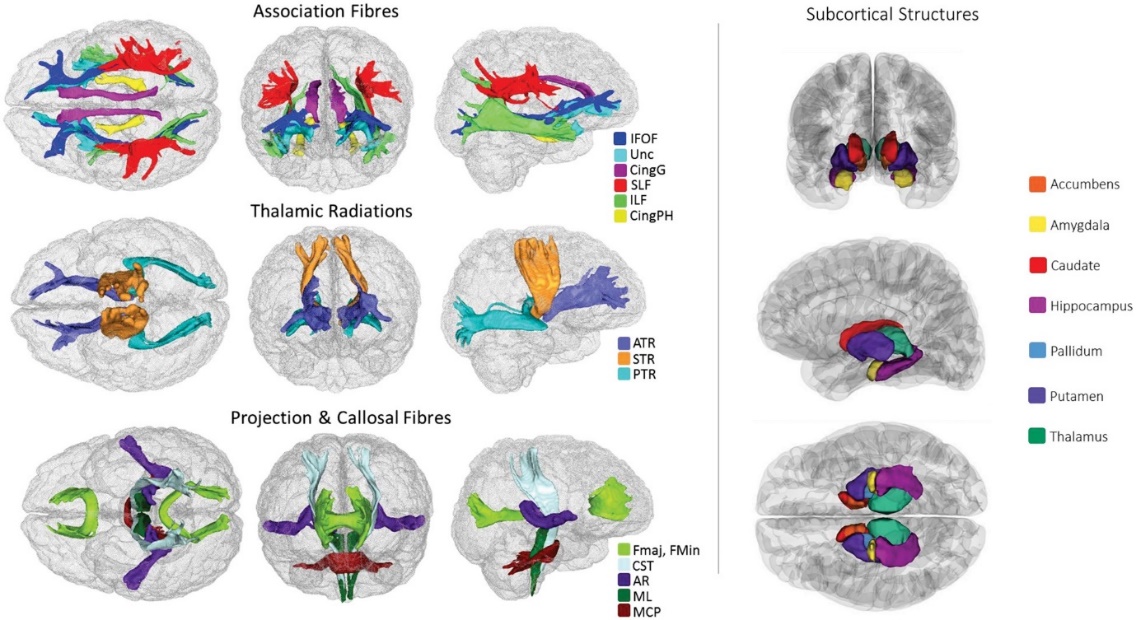

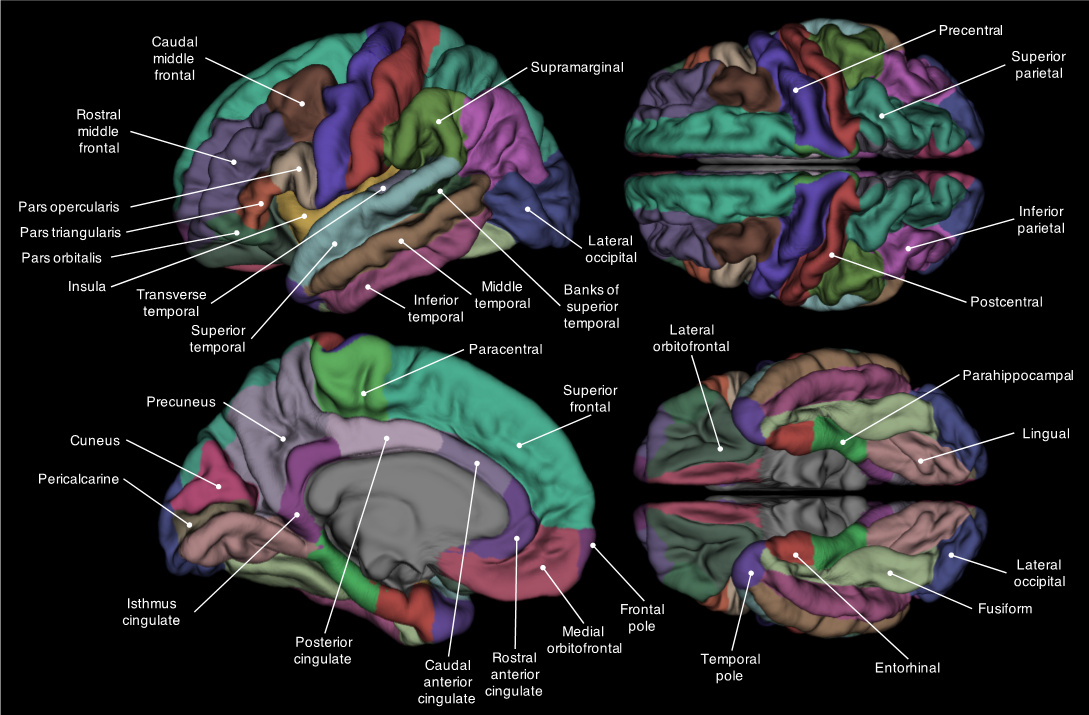

*Note*: AR, acoustic radiation; ATR, anterior thalamic radiation; Cing, cingulum (gyrus and parahippocampal); CST, corticospinal tract; Fmaj and Fmin (forceps major and minor); IFOF, inferior fronto-occipital fasciculus; ILF, inferior longitudinal fasciculus; MCP, middle cerebellar peduncle; ML, medial lemniscus; PTR, posterior thalamic radiation; SLF, superior longitudinal fasciculus; STR, superior thalamic radiation; Unc, uncinate fasciculus.

**Supplementary Table 4**: Anticholinergics scales identified in the present study. We considered anticholinergic scales that were available as complete lists of drugs, scored each drug for its anticholinergic potency, and did not require information on dosage. Grey shading indicates that the scale was not considered for further analysis. For two scales^3,4^, updated versions (Aging Brain Care, 2012; Carnahan, 2014, personal communication on 21.10.2019) were used. One scale^5^ was updated to include newer drugs from the UK market as has been done before^6^. The table was modified from a previous study^7^.

| **Surname of first author** | **Scale name** | **Year of publication** | **Reason for exclusion** |
| --- | --- | --- | --- |
| Summers^8^ | Drug Risk Number (DRN) | 1978 | Outdated (based on the date of publication and on new scales developed on its basis). |
| Han^9^ | Clinician-rated Anticholinergic Scale (CrAS) | 2001 |  |
| Aizenberg^10^ | Anticholinergic Burden Score (ABS) | 2002 | Publicly unavailable and no response from lead author to two email requests within a year. |
| Minzenberg^11^ | n.a. | 2004 | Based on a reference compound. |
| Ancelin^12^ | Anticholinergic Burden Classification (ABC) scale | 2006 |  |
| Carnahan^4^ | Anticholinergic Drug Scale (ADS) | 2006 (2014) |  |
| Hilmer^13^ | Drug Burden Index (DBI) | 2007 | Required information on drug dosage. |
| Chew^14^ | Anticholinergic Activity Scale (AAS) | 2008 |  |
| Cancelli^15^ | n.a. | 2008 |  |
| Rudolph^5^ | Anticholinergic Risk Scale (ARS) | 2008 (2013) |  |
| Ehrt^16^ | Revised Anticholinergic Activity Scale (AAS-r) | 2010 |  |
| Sittironnarit^17^ | Anticholinergic Loading Scale (ALS) | 2011 |  |
| Boustani^3^ | Anticholinergic Cognitive Burden (ACB) | 2008 (2012) |  |
| Whalley^18^ | n.a. | 2012 | Unavailable in full. |
| Durán^19^ | n.a. | 2013 |  |
| Faure^20^ | Drug Burden Index, International Version (DBI-WHO) | 2014 | Required information on drug dosage. |
| Klamer^21^ | MARANTE | 2017 | Required information on drug dosage. |
| Bishara^22^ | Anticholinergic effect on cognition (AEC) scale | 2017 |  |
| Briet^23^ | Anticholinergic impregnation scale | 2017 |  |
| Kiesel^24^ | n.a. | 2018 |  |
| Nery^25^ | Brazilian anticholinergic activity drug scale | 2019 |  |
| Jun^26^ | Korean Anticholinergic Burden Scale (KABS) | 2019 |  |

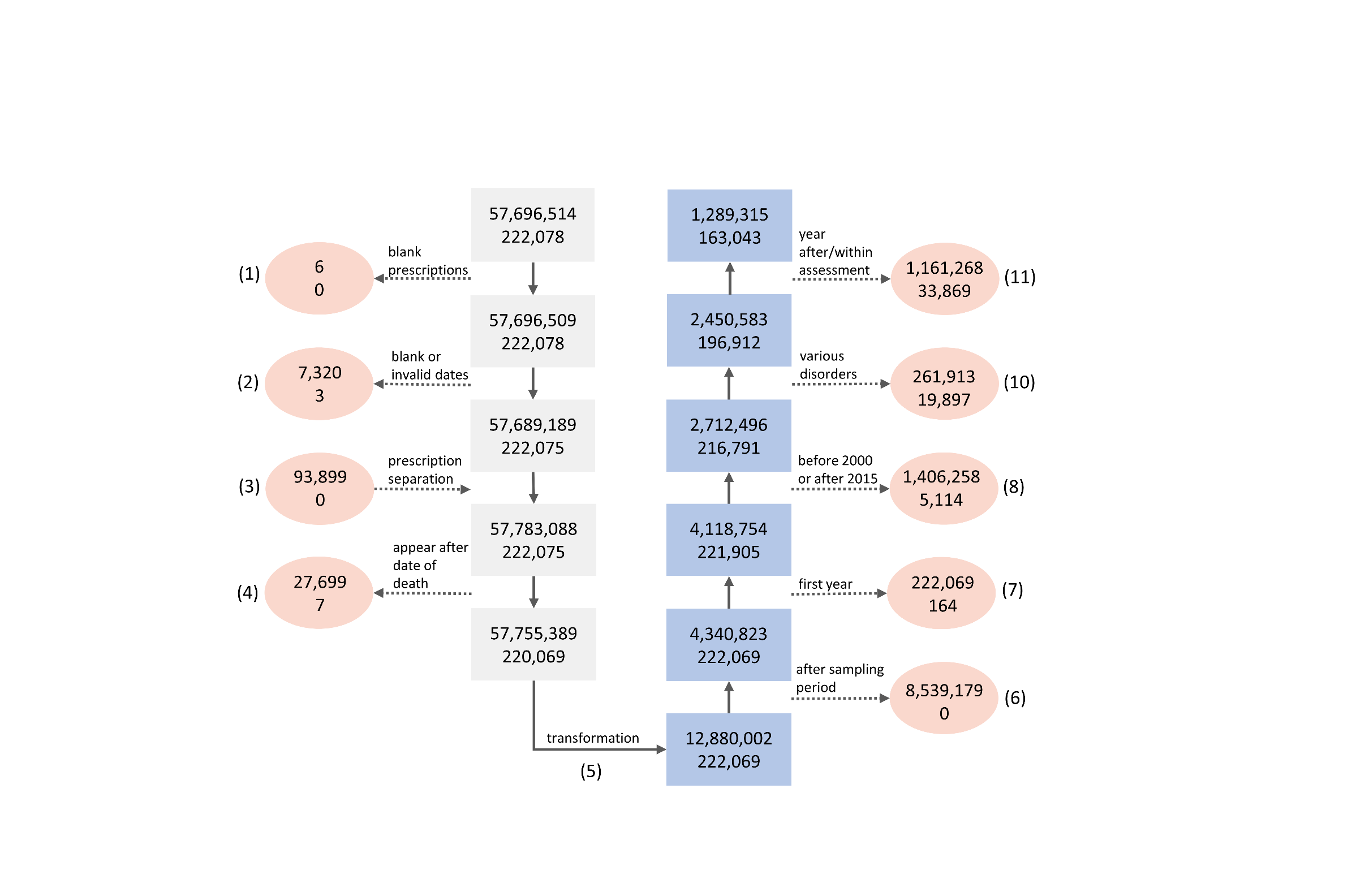
**Supplementary Figure 3**: The data-cleaning pipeline. The upper and lower rows of numbers represent the numbers of observation and participants, respectively. The rectangular boxes display the numbers of observations/participants at each point of the data-cleaning process, while the ovals display the numbers of observations/participants removed. The colour of the rectangles indicates the type of observation: in the grey boxes, the basic observation was a single prescription; in the blue boxes, it was a year-participant pair, with each pair representing the AChB of a single participant in a given year. The actions performed at each step are written above the arrows and signify the (1) removal of prescriptions without any content (i.e., no drug indicated), (2) removal of prescriptions without dates or impossible dates (e.g., in the future or far in the past), (3) separation of combination drugs into individual compounds, (4) removal of prescriptions that appear after the death of the participant, (5) transformation into the year-participant format, (6) removal of observations (generated in step (5)) occurring after the end of the prescription-sampling period for any participant, (7) removal of observations for the first year in the dataset for each participant (as it is unlikely to be complete), (8) removal of observations prior to the year 2000 and after the year 2015, (9) removal of observations for participants diagnosed with a disorder that may affect cognitive or brain function, (10) removal of observations after or within the year of the UK Biobank assessment.

**Supplementary Figure 4**: Q-Q plots of theoretical quantiles (x-axis) vs. standardised residuals (y-axis) for some models used in our study. Only the examples of models exhibiting most extreme kurtosis are shown. The examples here are for the association between AChB according to Durán et al. (2013) ^19^ on the one hand and either *g* (1), reaction time (2), visual memory (3), volume of the right pastriangularis (4), volume of the left pallidum (5), or volume of the right hippocampus (6), on the other.

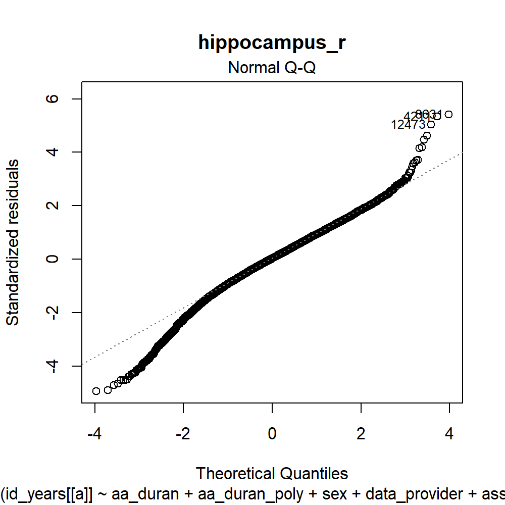

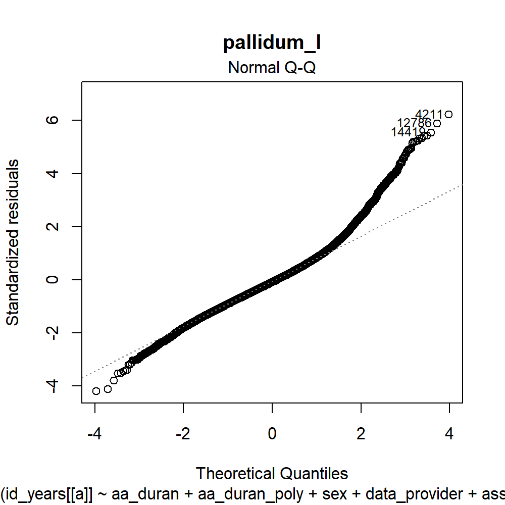

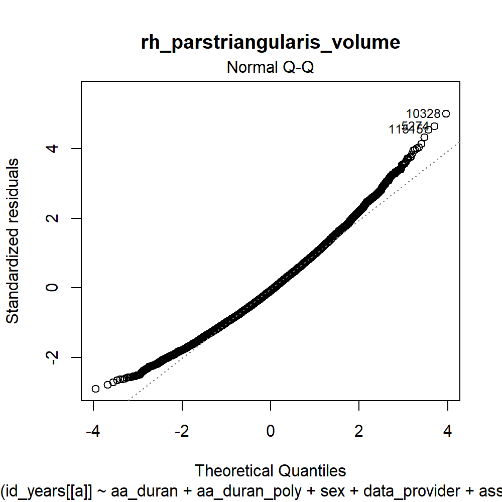

**Standardised residuals**

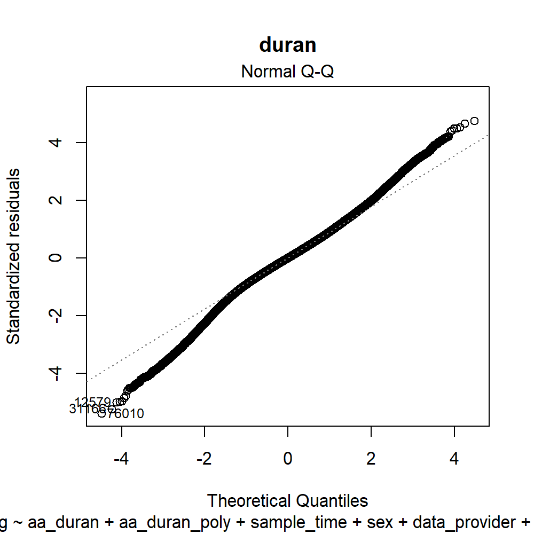

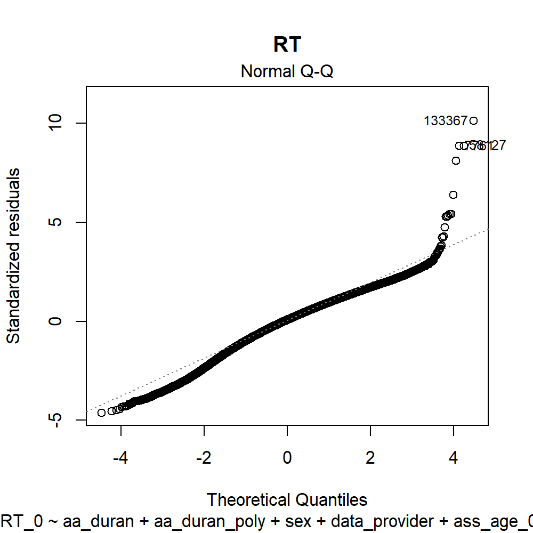

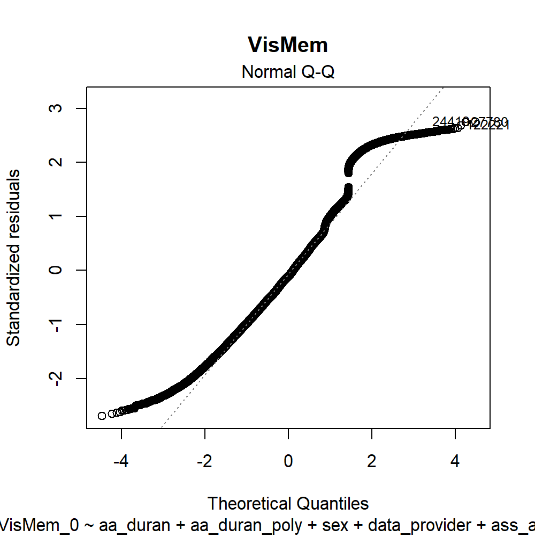

1

2

3

6

5

4

**Theoretical quantiles**

**Supplementary Figure 5:**
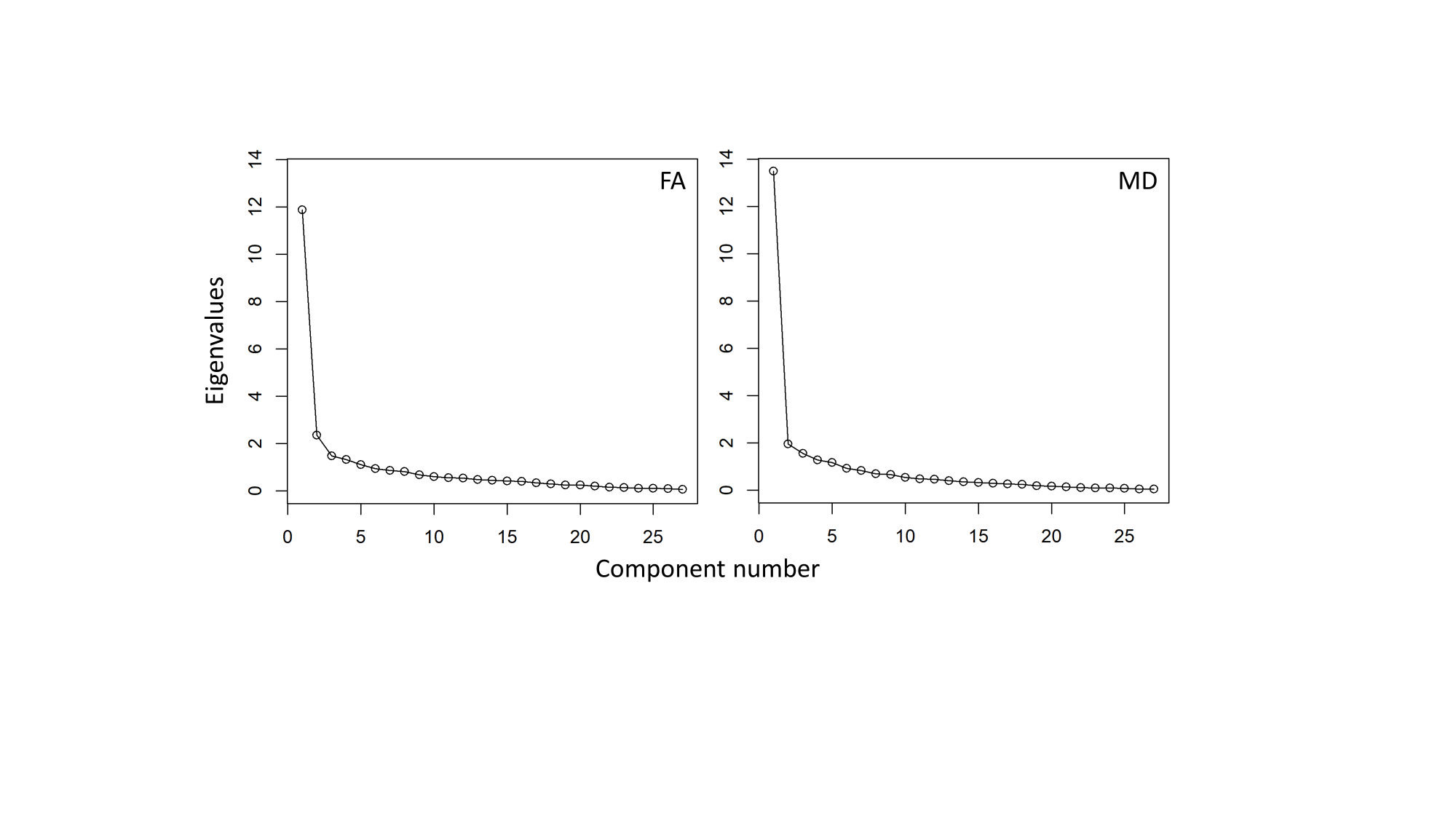
 Scree plots of eigenvalues for FA (**left**) and MD (**right**). The principal component for FA explained 43.9% of the total variance and the principal component for MD explained 49.9% of the total variance.

**Supplementary Text 1**: The models used in our study. (1.0) was used to compare anticholinergic scales (with each model using *g* as the outcome) and to compare cognitive tests (with each model using the scale by Durán et al. (2013) ^19^ as the predictor and a different cognitive test as the outcome). (1.1) is the basic polypharmacy model; it differs from (1.0) in that the number of non-anticholinergic drugs is the main predictor. (1.2) is the Polypharmacy Plus model that differs from (1.1) in that the number of anticholinergic drugs (according to any anticholinergic scale) is included as a covariate. (2.0) comprises of models that predict any measure of brain-MRI by AChB (according to the scale by Durán et al. (2013) ^19^ when not comparing scales) and include additional covariates. Polypharmacy models (analogous to 1.1 and 1.2, but with additional covariates as in 2.0) were also run for total brain volume when comparing anticholinergic scales.

$$1.0 Cognition_{scale}=lm(cognition \sim AChB_{scale}+non.anticholinergic.drug.number+age+number.prescription.years+data.provider+deprivation+smoking+alcohol+physical.activity+BMI+APOE+comorbidities+mood.disorder+anxiety.disorder+ schizophrenia+ diabetes+ hypercholesterolemia+hypertension+myocardial.infarction)$$

$$1.1 Cognition_{polypharmacy}=lm(g \sim drug.count+age+number.prescription.years+data.provider+deprivation+smoking+alcohol+physical.activity+BMI+APOE+comorbidities+mood.disorder+anxiety.disorder+ schizophrenia+ diabetes+ hypercholesterolemia+hypertension+myocardial.infarction)$$

$$1.2 Cognition_{polypharmacy.plus}=lm(g \sim drug.count+anticholinergic.drug.number++age+number.prescription.years+data.provider+deprivation+smoking+alcohol+physical.activity+BMI+APOE+comorbidities+mood.disorder+anxiety.disorder+ schizophrenia+ diabetes+ hypercholesterolemia+hypertension+myocardial.infarction)$$

$$2.MRI_{scale}=lm(MRI \sim AChB_{scale}+non.anticholinergic.drug.number++age+number.prescription.years+data.provider+deprivation+smoking+alcohol+physical.activity+BMI+APOE+comorbidities+mood.disorder+anxiety.disorder+ schizophrenia+ diabetes+ hypercholesterolemia+hypertension+myocardial.infarction+ age^{2}+age*sex+age^{2}*sex+head.position+ethnicity+asessment.centre)$$

**Supplementary Table 6**: Demographic and lifestyle characteristics of separately the imaging subsample and the rest of the sample after the removal of outliers. The columns indicate the median and IQR (or n and % for categorical variables) and the number of missing observations. The variables are not scaled. Note that the counts may not always add up to those depicted in **Table 1**, as the imaging subsample underwent separate data-cleaning before running the analyses.

| **Variable** | **Level** | **Imaging subsample** | | **Rest of sample** | |
| --- | --- | --- | --- | --- | --- |
|  |  | **Median (IQR) or n (%)** | **N missing** | **Median (IQR) or n (%)** | **N missing** |
| Age |  | 64.8 (11.9) |  | 58.6 (12.9) |  |
| Sex | Male | 8,072 (46.6) 9,265 (53.4) |  | 62,640 (43.8) 80,412 (56.2) |  |
| Deprivation |  | -2.7 (3.2) | 21 | -2.2 (3.9) | 149 |
| Alcohol consumption | Daily or almost daily Three or four times a week Once or twice a week Once to three times a month Only special occasions Never | 2,926 (17.0) 4,927 (28.6)  4,536 (26.3)  1,994 (11.6)  1,713 (9.9)  1,124 (6.5) | 117 | 27,778 (19.5) 32,812 (23.0)  38,103 (26.7)  16,225 (11.4)  16,430 (11.5)  11,388 (8.0) | 316 |
| Smoking | Current smoker  Previous smoker  Non-smoker | 561 (3.3) 5,744 (33.4) 10,867 (63.3) | 165 | 14,819 (10.4) 49,029 (34.5) 78,464 (55.1) | 740 |
| Physical activity | Strenuous  Moderate  Light | 2,232 (13.3) 11,346 (67.4) 3,246 (19.3) | 513 | 13,583 (10.2) 84,918 (63.8) 34,521 (26.0) | 10,030 |
| BMI |  | 25.8 (5.3) | 590 | 26.9 (5.8) | 987 |
| Region | England (Vision) Scotland England (TPP) Wales | 1,524 (8.8) 1,500 (8.7) 14,133 (81.5) 180 (1.0) |  | 12,592 (8.8) 7,862 (5.5) 105,849 (74.0) 16,749 (11.7) |  |
| Mood disorder |  | 2,126 (12.3) |  | 21,431 (15.0) |  |
| Anxiety disorder |  | 1,250 (7.2) |  | 14,037 (9.8) |  |
| Schizophrenia |  | 20 (0.12) |  | 555 (0.4) |  |
| Myocardial infarction |  | 370 (2.1) |  | 6,762 (4.7) |  |
| Diabetes |  | 766 (4.4) |  | 13,425 (9.4) |  |
| Hypercholesterolemia |  | 2,041 (11.8) |  | 27,259 (19.1) |  |
| Hypertension |  | 3,729 (21.5) |  | 49,191 (34.4) |  |
| Number of prior comorbidities |  | 135 (103) | 2 | 81 (92) | 47 |
| Polypharmacy |  | 49 (132) |  | 33 (92) |  |
| *APOE* carrier | ε2  ε3  ε4 | 2,224 (13.1) 10,498 (61.7) 4,286 (25.2) | 329 | 18,010 (12.9) 85,949 (61.6) 35,603 (25.5) | 3,490 |
| Ethnicity | British  Irish  Any other white background  White and black Caribbean  White and black African  White and Asian  Any other mixed background  Indian  Pakistani  Bangladeshi  Any other Asian background  Caribbean  African  Any other black background | 15,942 (93.0) 376 (2.2) 481 (2.8)  16 (0.09)  7 (0.04)  29 (0.17) 22 (0.13)  131 (0.76) 33 (0.19) 2 (0.01) 27 (0.16)  46 (0.27) 35 (0.20) 1 (0.006) | 189 | 128,246 (91.1)  3,307 (2.3)  3,906 (2.8)  169 (0.12)  94 (0.07)  204 (0.15)  250 (0.18)  2,006 (1.4)  606 (0.43)  47 (0.03)  434 (0.31)  1,005 (0.71)  538 (0.38)  25 (0.02) | 2,215 |
| Imaging assessment centre | Cheadle  Reading  Newcastle Bristol | 11,534 (60.7) 1,208 (7.0) 5,581 (32.2) 14 (0.08) |  |  |  |

| **Anticholinergic scale** | **N (%)** | **>1 n (%)** |
| --- | --- | --- |
| Ancelin et al. (2006) | 632,410 (4.3) | 19,656 (12.1) |
| Bishara et al. (2017) | 1,328,952 (8.9) | 41,871 (25.7) |
| Boustani et al. (2008*) | 2,158,487 (14.5) | 54,480 (33.4) |
| Briet et al. (2017) | 2,814,146 (18.9) | 60,975 (37.4) |
| Cancelli et al. (2008) | 778,080 (5.2) | 18,445 (11.3) |
| Carnahan et al. (2006*) | 1,642,406 (11.1) | 48,411 (29.7) |
| Chew et al. (2008) | 1,912,596 (12.9) | 58,773 (36.0) |
| Durán et al. (2013) | 3,118,524 (21.0) | 66,289 (40.7) |
| Ehrt et al. (2010) | 1,336,023 (9.0) | 37,931 (23.3) |
| Han et al. (2001) | 1,952,053 (13.1) | 54,311 (33.3) |
| Jun et al. (2019) | 2,075,559 (14.0) | 55,657 (34.1) |
| Kiesel et al. (2018) | 3,579,841 (24.1) | 62,889 (38.6) |
| Nery et al. (2019) | 2,745,039 (18.5) | 59,915 (36.7) |
| Rudolph et al. (2008*) | 1,108,629 (7.5) | 41,556 (25.5) |
| Sittironnarit et al. (2011) | 2,169,407 (14.6) | 55,948 (34.3) |

**Supplementary Table 7**: Numbers (and % of total prescriptions) of anticholinergic prescriptions per anticholinergic scale and numbers (and % of total sample) of participants prescribed at least one anticholinergic prescription in the sampling period. The data include the period between the year 2000 and the year prior to attending the assessment visit (differs between participants).

**Note*: some scales were updated after their initial date of publication, as noted in **Supplementary Table 4**.

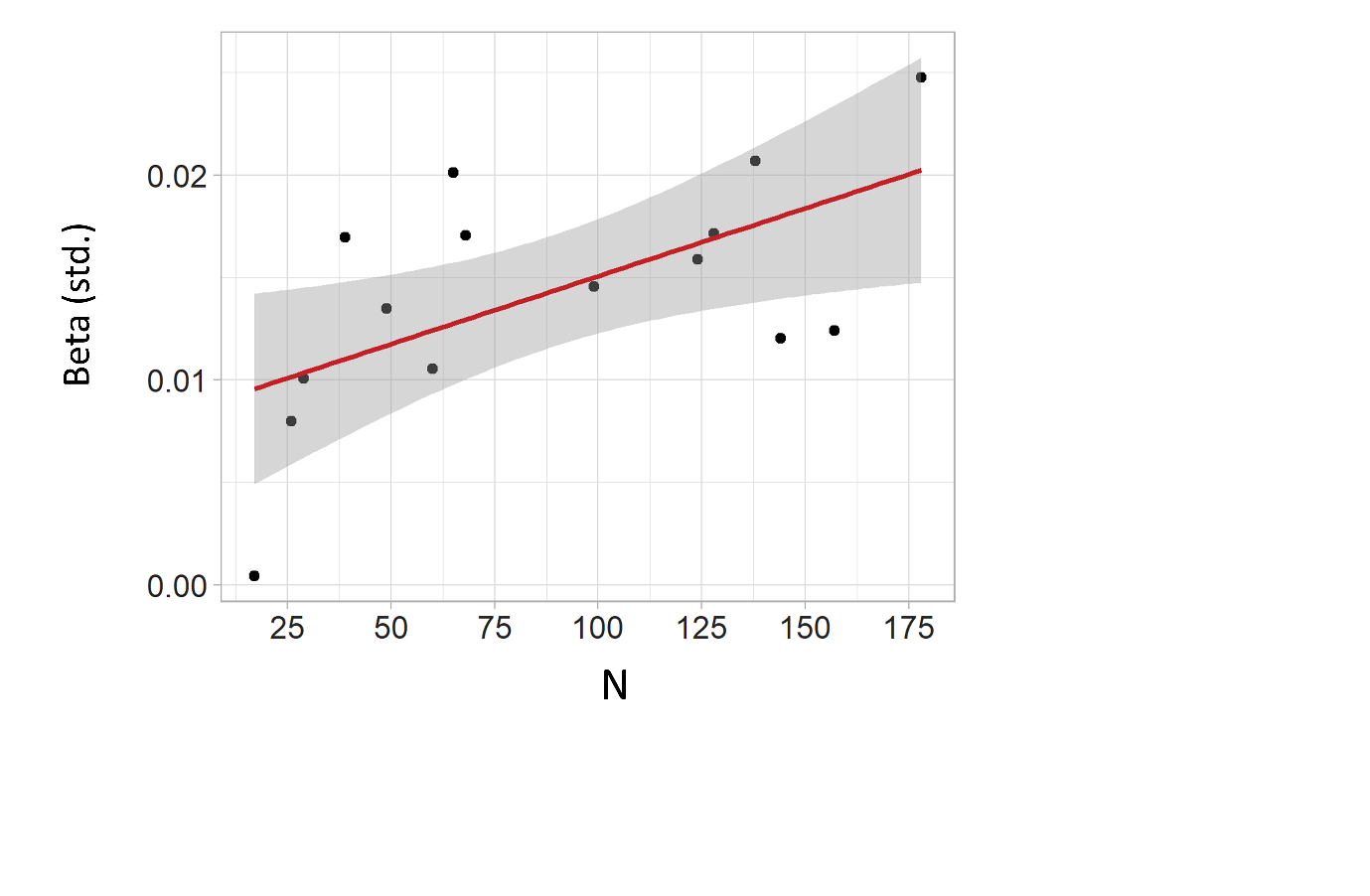
**Supplementary Figure 6:** Scatterplot for the association between the number of drugs identified as having an anticholinergic effect and the association with lower cognitive ability. The x-axis represents the number of drugs identified as possessing anticholinergic effects, the y-axis represents the absolute value of the standardised β for the association between AChB and *g*. Each dot in the scatterplot represents an anticholinergic scale. The red line represents the line of best fit, with grey shading indicating the 95% CI.
